# Connection between two historical tuberculosis outbreak sites in Japan, Honshu, due to a new ancestral L2 sublineage

**DOI:** 10.1101/2021.06.29.21259634

**Authors:** Christophe Guyeux, Gaetan Senelle, Guislaine Refrégier, Emmanuelle Cambau, Christophe Sola

## Abstract

By gathering an initial collection of 680 public Sequence Read Archives from the *Mycobacterium tuberculosis* complex (MTC) including 190 belonging to the lineage 2 *Beijing*, and using a new bioinformatical pipeline, *TB-Annotator*, that analyzes more than 50,000 characters, we characterized a phylogenetically significant L2 sublineage from isolates found in Tochigi province, that we designate as *Asia Ancestral 5*. These isolates harbor a number of specific criteria (42 specific SNPs) and their intra-cluster pairwise distance suggested historical and not epidemiological transmission. These isolates harbor a mutation in *rpoC*, and do not fulfill, either the *Modern Beijing* lineage criteria (L2.2.1.2.2) or the other currently known *Ancestral Beijing* lineages described so far. *Asia Ancestral 5* isolates do not possess *mutT2* 58 and *ogt* 12 characteristics of *Modern Beijing*, but possess ancient evolutionary characteristics. By looking into the literature, we found in a reference isolate ID381, found in Kobe and Osaka, and defined as “G3”, 36 out of 42 shared and specific SNPs in the same sublineage. Using this study, we also confirmed the intermediate position of the *Asia Ancestral 4* recently described in Thailand and suggest an improved classification of the L2 that now includes *Asia Ancestral 4 and 5*. Increasing the recruitment to around 3000 genomes (including 642 belonging to L2) confirmed our results and suggest new historical ancestral L2 phylogenetically relevant branches that remain to be investigated in detail. We discuss some anthropological and historical data from Japan history and its link to Korea and China. This study shows that the reconstruction of the early history of tuberculosis pandemia in Asia is likely to reveal complex patterns since its emergence.

## Introduction

With 10 millions new cases in 2020, and around 500,000 Multi-drug resistant cases, Tuberculosis (TB) is far from being eradicated [1]. There are nine recognized genetic lineages in the bacterial pathogen agent, *Mycobacterium tuberculosis* complex (MTC), and one of these, lineage 2 (L2), is of particular interest due to the emergence of multi-drug-resistance tuberculosis (MDR-TB) and extremely drug-resistant tuberculosis (XDR-TB), that were described in numerous studies run in Russia, Europe, Africa, Asia and America [2-6]. L2 finds its origin in China and is predominant in Asia [7, 8]. The adaptation ability of these bacilli to develop specific pathogenic features including compensatory mutations in genes that restore their fitness have largely contributed to their epidemic success and to the global spreading of multidrug resistant strains [6, 9-11].

The epidemic bursts of L2 in the 90s can be linked to contemporary historical and geopolitical events, such as: (1) the fall of the former USSR, (2) the poor individualized TB treatment and follow-up of prisoners in Former Soviet Union (FSU) republics, that likely contributed when freed, to the seeding of the general population in Russia with L2 isolates that had become multidrug resistant in prisoners populations, (3) the increasing global trade share of China in the world, and in particular in Middle-East, South and East-Africa, and more generally around the African coasts, (4) the current Covid-19 pandemia with its dramatic consequences on global health [12-15].

The discovery of the *Beijing* Lineage can be dated back to 1995, even if the existence of the “W” strain, a member of L2, was known since its discovery in New York in the early 90s [16, 17]. It was first characterized by its high IS*6110* Insertion Sequence copy number (16 to 20 copies), with a diagnostic copy in the *dnaA-dnaN* also designated as the *NTF* region [18, 19]. No lineage such as L2 has been fostering so many genetic, genomic, molecular epidemiological and public health studies, whether for detection of multidrug resistance (a feature rapidly shown to be associated to L2), for improvement of its taxonomy through deletion region analysis, for a better understanding of its genetic population structure, and of course for its surveillance [4, 20, 21]. This was also made possible thanks to Variable Number of Tandem Repeats (VNTR) analysis, by Regions of Difference (RD) analysis using DNA hybridization, and lastly by the Whole-Genome Sequencing approach that led to single nucleotide polymorphisms (SNP) discoveries [22, 23]. Very large outbreaks inside this lineage were shown to have independently emerged in Russia, in Europe, in Central Asia, in South Africa, and of course in China [24].

L2 were previously classified as “*typical*” (modern) or “*atypical*” (ancestral/ancient) based on IS6*110*-RFLP patterns and later on Deletion Region analysis [25, 26]. Today, the L2 lineage is split into two main sublineages, L2.1 and L2.2. L2.1 is known as “Proto-Beijing” and is found mainly in Southern China, particularly in Guangxi province [27]. Among the major characteristics is the fact that it possesses the L2 characterizing RD105 deletion but not RD207 [28]. Recently a rare L2.1 clone was shown to have become ultra-drug resistant [28]. Such findings raise the issue of genetic background on adaptation capacity and acquisition of multi-drug resistance. L2.2, or the “Beijing” strain, as defined by spoligotyping SIT1 most of the time, is composed of several sublineages broadly categorized into the “*Ancestral*” and “*Modern*” Beijing strains. L2.2.2 defines the Asia Ancestral 1 group, whereas L2.2.1 defines all others, and that do not fit anymore with the “modern Beijing” designation [29]. The switch from “*Ancestral”* to “*Modern”* Beijing would be associated to the presence of at least one IS*6110* copy in the NTF and the presence of the MutT2 G->C mutation in position 1286766 and *ogt* codon 12 (C1477596T, ogt12) [9, 30, 31]. The “*Ancestral Beijing*” strains form a cluster made up, until now, of three “*Asia Ancestral”* lineages (Asia Ancestral 1 to Asia Ancestral 3, AAnc1 to AAnc3) [31], to which a fourth “*Asia Ancestral 4*” (AAnc4), found in Northern Thailand, was recently added [32]. RD181 is not deleted in L2.1 and L2.2.2 whereas is deleted in all other sublineages [31].

The “*Modern*” Beijing represents the clade that is at the end of the phylogenetic tree and it encompasses at least 7 large and epidemiologically acknowledged sublineages (Asian African 1, Asian African 3, Europe/Russia B0/W148 outbreak, Central Asia, Asian African 2, Asian African2/RD142, Pacific RD150) [31]. Interestingly, apart from the SNPs discovered these recent years, other major genomic rearrangements were shown to have happened in some L2 genomes [33, 34]. Modern Beijing strains are responsible of most but not of all of the recent MDR-TB outbreaks and are more transmissible [4, 35, 36].

However, apart from these well MDR-TB characterized strains and lineages, there remain many L2 isolates that do not fit to any known sublineages [37]. As an example, 26 Clonal Complexes (Bmyc1 to Bmyc26) were described based on 3R gene polymorphisms, however some of these types (e.g. Bmyc6 or Bmyc26) remained unclusterized based on the most recent classification scheme [31]. This suggests that some improvements in the Shitikov L2 classification scheme are possible; meanwhile other L2 sublineages were described such as in Thailand or Japan, and their history should also be linked to the global L2 history by performing comparative genomics [6, 23, 28, 32, 38-42].

The difficulties to define L2 epidemiological clusters using IS*6110*-RFLP, the impossibility to study this lineage by spoligotyping (almost all strains harbor the SIT1 spoligotype with only spacers 34-43 left) promoted the use of deletion regions and MIRU-VNTR to decipher its complex population structure [43]. One example of these difficulties was the need to add hypervariable VNTR Loci for L2 molecular epidemiological studies [44, 45]. A 15-9 VNTR signature designation was also introduced to facilitate designation of MTC epidemiological clones [45]. Hence, the introduction of Whole Genome Sequencing (WGS) and the building of very large databases to create representative worldwide collections is the best way to develop a more comprehensive and precise knowledge of this complex lineage [4, 23, 46]. The Merker *et al*. study represents the greatest endeavour to date to get an *in-depth* characterization of the L2 lineage [23]. It showed the existence of 6 clonal complex (CC1 to CC6) and a basal sublineage (BL7) [23].

RD and SNP-based phylogeny provides the best tools to classify L2 isolates. However, an even more refined evolutionary scheme, can be based on IS*6110* insertion sites that allows a fine study on microevolution of all lineages [47, 48]. Indeed, the adaptation to an IS*6110* high copy number and diversity of insertion sites in L2 are likely to be linked to increased mutation rates and are likely to provide in some cases a higher fitness of some strains [36, 49]. The use of MIRU-VNTR results in association to WGS is also interesting but seems to reach some limits [39]. Recently, thanks to a very useful standardization of all previously identified sublineages naming, a very clear population structure of L2 emerged, with 11 main L2 sublineages [31].

The history of the evolution of L2 still contains many local and global shadow zones, in particular concerning the dating of the split between L2 and L3, and the precise dating of emergence and geographical origin of the various sublineages and on their specific phylodynamics [23, 30]. Hirsch *et al*. suggested that East Asian and Philippines human populations carrying *M. tuberculosis* belonging to distinct lineages may have split 240–1,000 years ago [50]. Merker *et al*. generated estimates for the time to the most recent common ancestor with a TMRCAs of ∼6,000 and 5,000 years for CC6 and BL7 (the most ancient estimated by coalescence), and a TMRCA of ∼1,500 years for CC5 (the most recently appeared) [23]. More recently however, different estimated emergence dates were given [8]. In this study, an estimate of 2200 Before Present (BP) was given for the coalescence between Proto-Beijing and ancestral Beijing, and an estimate of 1300 BP for the split between all “ancestral-Beijing” lineages [8]. 900 BP would correspond to “Proto-Beijing” expansion whereas expansion of “Modern Beijing” would have appeared only around 500 years ago. The place of Japan, and its relation to China was also raised by some authors, however no precise emergence date was suggested [51]. Hence, as seen above, there remain many uncertainties on the exact dating of origin, and various split and bursts of L2.

The geographic origin of the emergence of L2 is almost as blurry as its dating. According to some authors, north-central and northeast China could have been the initial spreading center [51]. According to others, based on large differences in the prevalence of the ancient L2 lineages in China and on the higher genetic diversity observed in the south-west province of Guizhou, South-China would be the geographical origin of L2 [27, 30]. Indeed, Guizhou counts 17 ethnical minorities, and most of the non-officially acknowledged ethnic groups of China are located in this province. South-East-Asia shows a higher human genetic diversity than the north-east-Asia [52]. Arguing in favour of a South-China origin, a recent paper described the existence of an “*Asia-Ancestral 4*” L2 sublineage, that was found in the north of Thailand in a city named Chiangrai [32]. Chiangrai has been inhabited since the 7th century and peopled by tribes now residing in Thailand, Myanmar and Lao [32]. In this place, Ancestral Beijing strains are associated with ethnic minorities, Akha and Lahu tribes, who mostly migrated from Southern China [32].

In Japan, one of the main characteristics of the tuberculosis global history is the presence of a high and still poorly characterized genetic diversity of Ancestral L2 Beijing strains [23,42,44]. Mokrousov had detected that MIRU-VNTR diversity was quite important and specific in Japan [51]. Indeed, contrarily to what is observed in many other countries, L2 outbreaks in Japan are often due to ancestral Beijing strains, especially in aged people, that were not vaccinated by BCG [39, 42, 44, 53]. Hence, tuberculosis in Japan could be as ancient as in China or almost. Indeed, peopling of Japan is known to have originated through the Korean peninsula very early in the history of human migrations, followed by what is known as the Jomon period [54]. During the following Yayoi period, technology and agriculture was imported from Korea and from China [54].

We are developing the “*TB-Annotator*” project, a new bioinformatical pipeline whose aim is to perform data-mining of MTBC genomic diversity for both evolutionary biology and epidemiology using most of the information contained in Sequence Read Archives (SRAs). During the advancement of this long-term research project, we studied a set of L2 isolates from a prefecture located in Central Honshu in Japan [42]. In this study, 169 clinical isolates of *M. tuberculosis* were obtained from as many TB patients residing in Tochigi Prefecture [42]. Patient strains were collected in 2007 or in 2013 and were more likely to belong to the Ancestral (Atypical) L2 Lineage (n=79) than to the Modern (typical) (n=32) (see also Table 1 in [42]). Three patients only, out of the 79 bearing an Atypical Beijing strains were not born in Japan [42]. Japanese patients suffering from an Atypical Beijing were more likely to be found in the Central part of the Tochigi prefecture [42]. All the isolates had been characterized by WGS and analyzed using a bioinformatical pipeline, coined as “CAST” [55], however an *in-depth* comparative study of these isolates with other ancestral L2 had not been performed and we thus decided to study these genomes in more detail using our pipeline.

As more and more MTC genomes from isolated population are described, i.e. mountainous areas or isolated ethnic tribes or islanders, it becomes more and more likely to find rare though interesting L2 isolates that have their own specific history [32]. We describe in this article such a finding, i.e. the characterization of a new L2 sublineage from Japan and that is exclusive from Japan for the time-being. We also confirm the “*intermediate*” (i.e. between ancestral and modern) character of the AAnc4 sublineage recently described at the light of this comparative genomic analyses, and link these two lineages to the latest public and unified L2 classification scheme [31].

## Material and Methods

### Selected genomes

We downloaded a set of 680 SRAs; these genomes were selected to include the maximum of diversity of the currently described MTC sublineages characterized so far, including recent landmark papers [56, 57]. The list of these SRAs is shown in **Supplementary Table1** and includes 190 L2-assigned SRAs of all sublineages except for the terminal Pacific RD150 sublineage. From [32], we selected 28 SRAs labeled as “Asia ancestral 4”. From [42], we initially included a total of 158 representative genomes, however we discarded the SRAs for which the coverage was either too weak or for which it was impossible to rebuild the spoligotype using CRISPR-builder-TB [58, 59].

### Brief description of the *TB-Annotator* pipeline

The full version of *TB-Annotator* is going to be released in another article. In brief, regarding the set-up of the platform, the processes are the following: SRAs of interest are selected and kept only provided a certain number of conditions that together reinforce the reliability of the data: they have read length>75bp, if clean reads file is at least 100 Mo, and if CRISPR could be reconstructed using CRISPRbuilder-TB [59]. For each SRA, in addition to reconstructing the CRISPR-Cas region using CRISPRbuilder-TB and apart from collecting NCBI information on the genomes, two scripts successively: (1) search for SNPs according to reference catalogues (**Supplementary Table2**) totaling more than 50,000 SNPs, including drug-resistance related SNPs, phylogenetic SNPs as per 26 studies contributing to SNP-based classification ([23, 29, 31, 56, 57, 60-80]; (2) search for additional SNPs in each new isolate based on H37Rv reference sequence; (3) look for the presence/absence of H37Rv genes as annotated in mycobrowser (https://mycobrowser.epfl.ch/), (4) look for the presence/absence of deletion regions; (5) identify insertion sites of all known Insertion sequences in MTBC. The CRISPR locus is rebuilt using a dedicated and previously published script (43/68/360 Spacers format) with assignment of a Spoligo-International-Type (SIT) tag; the application produces an ordered list of spacers/repeat with variants and IS*6110* insertion sequences if present [58, 59, 81]. Final scripts allow to produce a phylogeny based on the list of studied characters and using RAxML and SplitsTree [82, 83].

All computations have been performed on the “*Mésocentre de Franche-Comté*” supercomputer facilities (141 nodes, 2292 cores, 9,27 To memory, 74,2 CPU Power TFlops, 66,4 GPU TFlops), using adequate commands. Apart from the Excel results file, the final phylogenetic tree on 680 selected genomes was built using SplitsTree to provide a graphical display of the results and using proprietary python scripts allowing queries to be made [83].

### SNP-based classification of L2 sublineages

In order to first assign the 680 selected SRAs into known lineages and sublineages, we used the reference list defined in **Supplementary Table2**. Full origin and classification results on the 190 SRA of the L2 isolates, as produced by “*TB-Annotator*” are found in the **Supplementary Table3**.

### Definition of an unclassified Asia Ancestral 5 sublineage (AAnc5)

After assessment of existing known L2 sublineages, the unknown branch (**Figure 1B**) was investigated in detail and was shown to originate only from Japan SRAs. The bioinformatical pipeline, based on its graphical user interface, allows to select and display new exclusive or shared SNPs and specific characteristics, that were further investigated.

**Figure 1.**
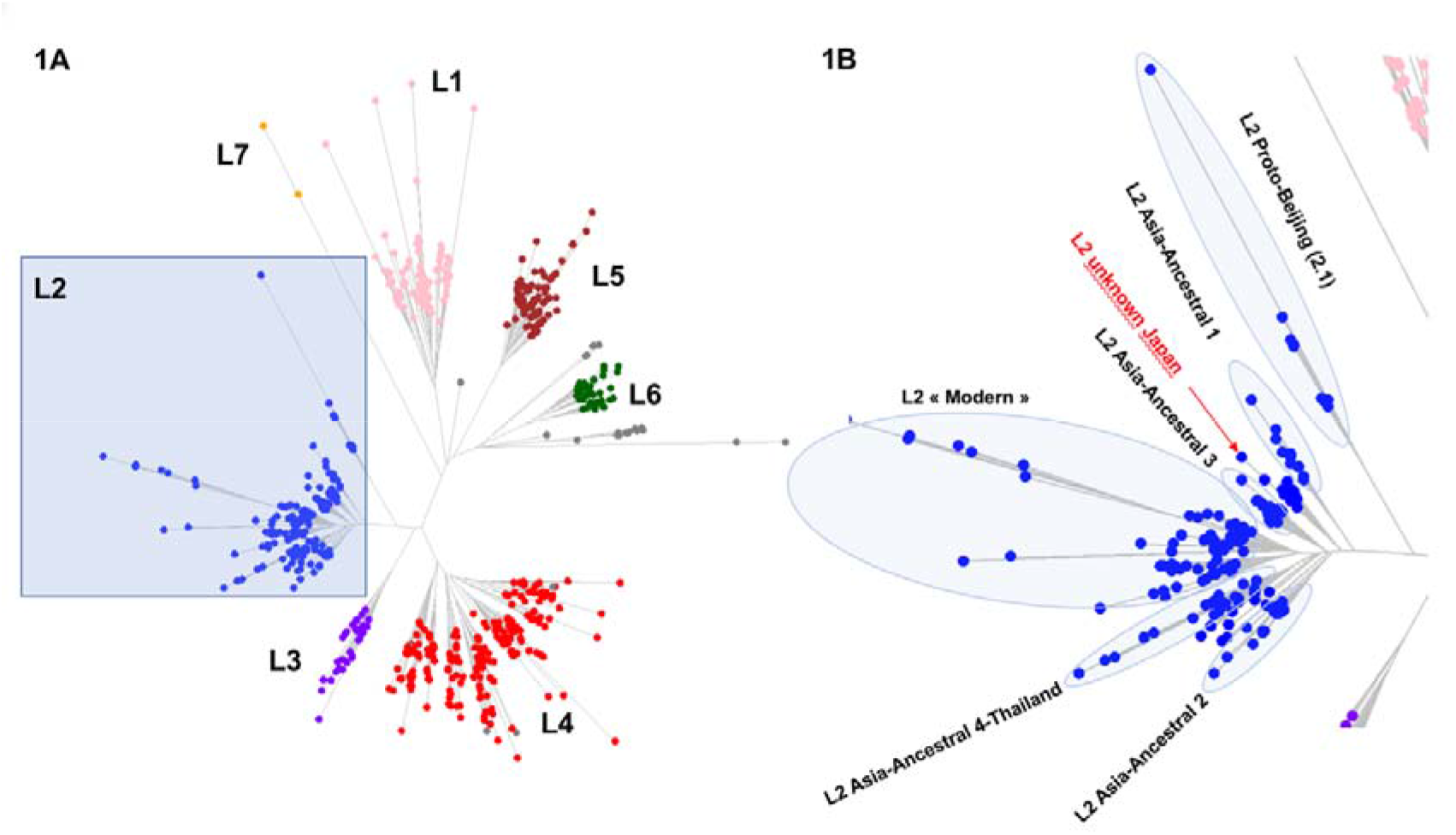
Left part (1A); *TB-annotator* unrooted phylogenetic tree on 680 SRA-derived data. L2 samples are shown in blue. Right part (1B); close-up on the Lineage 2 with all known branches named except in red the new unknown ancestral Japan sublineage we design as Asia Ancestral 5.

### Use of *TB-annotator* platform to explore diversity

The *TB-Annotator* interface is meant to explore diversity among TB samples. Presently, 680 samples representative of TB diversity were implemented. Other samples can be added upon request. The default layout presents the samples phylogeny. One specific function allows to identify the genetic markers that are specific to selected samples, either based on a graphic mode or on predefined filters. Useful metrics such as intra-cluster pairwise SNP distance can be computed. A second layout allows to identify the countries from which the selected samples were collected on a map. At the time of writing a new version with 3380 genomes confirmed our results.

## Results

### 1. In depth analysis of known SNPs phylogenetical markers within the 190 L2 genomes, discovery of a new L2 Sublineage

We implemented a representative set of samples from L2 and the 169 samples from the Tochigi province study, Honshu, onto the web platform *TB-annotator* [42]. Regarding the presence of phylogenetic SNPs and clustering in the phylogeny, as expected, all samples carrying the L2 SNP were found in the same phylogenetic branch (**Figure 1**), and 186 of the 190 isolates inside this branch carried this SNP (4 L2.1 isolates were missed). This branch can thus be considered as L2. Among these L2 samples, nine were Proto-Beijing (L2.1), ten were AAnc1 1 (L2.2.2) and the 171 isolates left were confirmed to fit the former L2.2.1 definition using Coll’s classification scheme [29]. This L2.2.1 sublineage, as previously mentioned, should not be referred to as “modern” Beijing since two of its sub-branches, AAnc 2 and 3 have been defined since [31]. These 171 L2.2.1 isolates shared a list of 31 exclusive SNPs (results not shown).

Going further in the analysis, 93 isolates from L2.2.1 were harboring the MutT2 G1286766C “modern Beijing” characteristic SNP. Out of these 93, 65 samples were also harboring the second “modern Beijing” signature, i.e. C1477596T SNP in *ogt* [32] while the 28 remaining harbored Asia Ancestral SNPs.

The “modern” Beijing (n=65), could be split into Asian African 1 (n=2), Asian African 3 (n=5), Asian African 2 (AAfr2, n=16), Asian African 2-RD142 (AAfr2-RD142, n=1), Central Asia (n=13), B0/W148 (n=2) and there were a remaining 26 “unclassified modern Beijing” isolates, that did not fit any described modern sublineage definition, and that were not investigated further in this study. No Pacific RD150 isolates was included in this study. All suggested binary names fit with Shitikov *et al*. scheme and we present a new improved classification that includes the recent discovery of “Asia Ancestral 4 and 5” (**Figure 2**).

**Figure 2:**
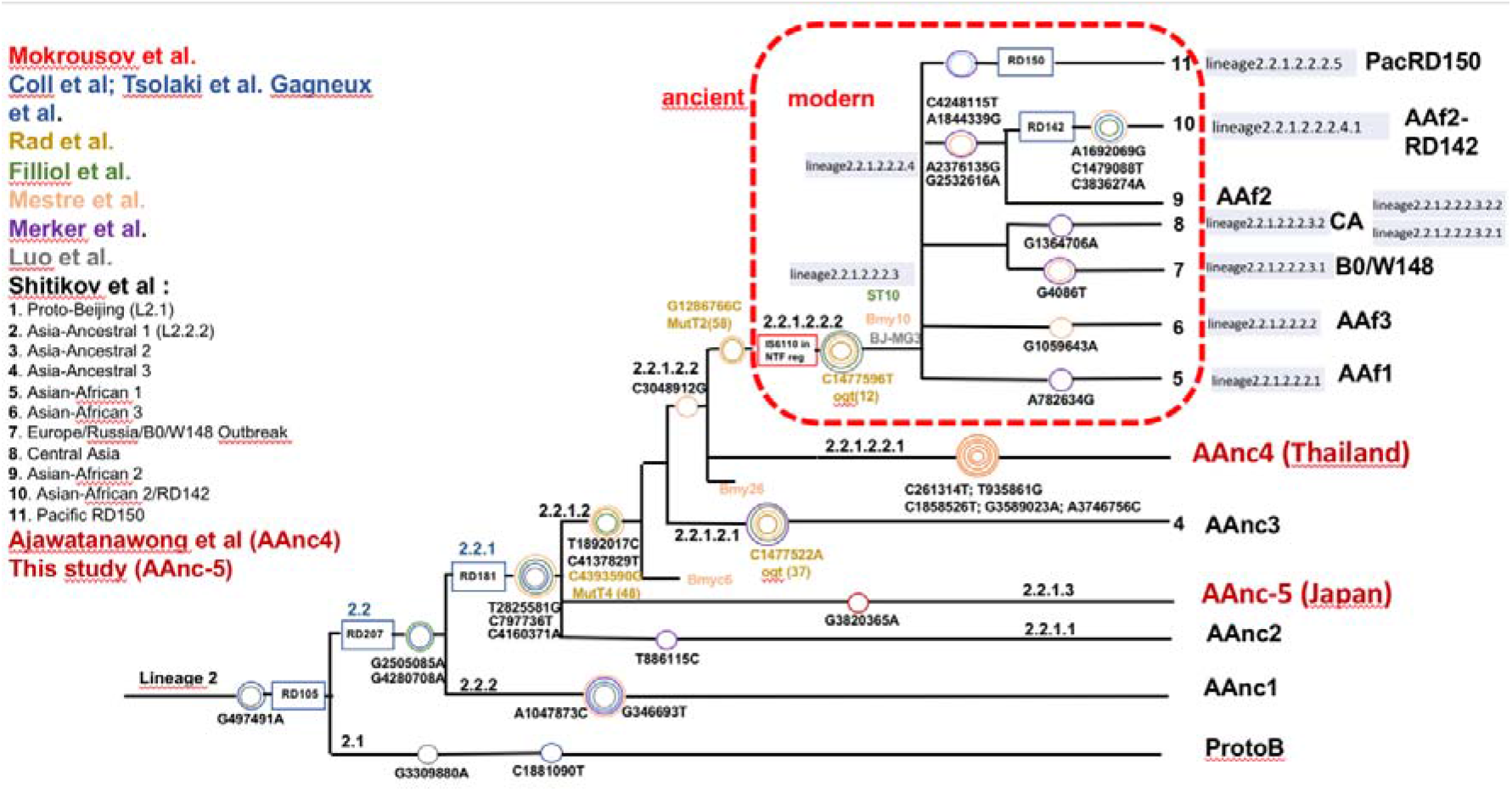
Mtb lineage 2 dendrogram representing the major phylogenetic groups and informative genetic markers (inspired and redrawn from Shitikov *et al*. 2017; note that on this new figure, SNP position of MutT2 and ogt 12 were corrected compared to the original figure where position were inverted). Bmyc6 and Bmyc26 were not considered in the binary naming suggested here.

As shown on **Figure 1**, the “ancestral” Beijing (n=124) could be split into Proto-Beijing (L2.1; n=9), Asia Ancestral 1 (L2.2.2-AAnc1, n=10), Asia-Ancestral 2 (L2.2.1.1-AAnc2, n=22), Asia-ancestral 3 (L2.2.1.2.1-AAnc3, n=9), “Asia-Ancestral 4” (L2.2.1.2.3*-AAnc4, n=27) and an unknown branch (suggested L2.2.1.3*-Asia Ancestral 5 or AAnc5, n=20); there still remains 27 unclustered and uncategorized ancestral Beijing.

The reanalysis of the phylogenetical SNP set described by Shitikov *et al*. confirmed all the phylogenetical SNPs specific of the Ancestral Beijing lineages up to the definition of the “modern” Beijing. The MutT2 SNP (G1286766C) is a good phylogenetic marker as it is present only in L2 modern sublineages whereas T1892017C, C4137829T and C4393590G (MutT4) are present in some ancestral sublineages. It is clear from the SNPs results and from the phylogenetical trees shown in **Figure 1** that the AAnc4 branch is intermediate, as suggested by their inventors, and is neither a *bona fide* modern L2 isolate nor a truly ancestral lineage [32]. Conversely, the 20 isolates that we studied from Japan, further designated as Asia Ancestral 5 (AAnc5), all fulfill the ancestral Beijing criteria since they do not possess the expected MutT2 (58) SNP nor any other characteristic of modern Beijing. They also branch before AAnc4. In all cases, the clear-cut split of L2 isolates that either possess none, one, or the two MutT2 (58) and ogt (12) SNPs confirms that these two markers are excellent phylogenetical ones. Based on the results obtained, we decided to adapt the classification scheme of Shitikov *et al*. to take into account all recently described sublineages (**Figure 2)**.

### 2. Description of the new “Asia-Ancestral 5” AAnc 5 Japan sublineage

#### 2.1 SNP-based results

We further characterized the Japan “Asia-Ancestral 5” (AAnc5) (n=20) that branches between AAnc 1 and 3 (**Figure 1**). These isolates can all be defined by many exclusively shared SNPs. The complete results matrix produced by the *TB-Annotator* pipeline on the 190 L2 genomes can be found in **Supplementary Table3**. In the future versions, reports will be downloadable from a dedicated website. The list of 42 exclusive SNPs, found in 16 of these 20 isolates is shown in **Supplementary Table4**. An interesting observation is that all the AAnc5 strains harbor a non-synonymous mutation in 765140 (G->C) in *rpoC*. This results will be discussed in detail further. Three other genomes (2 in other L2 sublineages and one in a L4 sublineage) also carry this *rpoC* mutation suggesting independent acquisition.

This new phylogenetical branch of L2 is believed to have emerged after the birth of AAnc1 but before the ones of AAnc2 and 3. A pairwise distance matrix between these isolates was constructed (**Supplementary Table5)**; the future pipeline will systematically compute the intra-branch SNP distance for clusters or selections of interest below 50 SRAs. The pairwise distance between these samples shows between a minimum of 166 SNPs (between DRR157280 and DRR157281) and a maximum of 439 SNPs distance (between DRR130203 and DRR034366) between AAnc5 isolates, thus excluding recent transmission (**Supplementary Table 5**). Assuming a 0.3 SNP mutation rate per year per genome, these strains might have diverged approximately between 250 to 600 years from their MRCA. Our results also confirmed the intermediate character of the “Asia Ancestral 4” (AAnc4) which was described in Chiangrai in Thailand [32]. If we accept the suggestion of timing of AAnc4 emergence or expansion around the 7^th^ century in Thailand, (start of Chiangrai), then AAnc5 could have been introduced into Japan earlier than this century, in line with archeological information [32] [84, 85]. A second list of 46 exclusively shared SNPs, found between two very distant isolates on a specific subbranch of AAnc5 (DRR034381 and DRR130203, pairwise distance: 417 SNP) AAnc5 is also shown in (**Supplementary Table5)**.

#### 2.2 *in Silico* spoligotyping and reconstruction of the CRISPR locus structure of AAnc 5 using *TB-Annotator*, IS*6110* copy number and insertion sites

One of the strengths of the *TB-Annotator* pipeline is that is also reconstructs the CRISPR by integration of the CRISPR-builder TB results [58, 59]. The 20 AAnc5 isolates showed 6 different spoligotype patterns, which was quite unexpected for an L2 sublineage (**Supplementary Table6**); most of these patterns have been previously described in the SITVIT database (SIT1, SIT190, SIT269, SIT1364, SIT1674), however one remained undefined as SIT”X”. No SNP variants were found in spacers and repeats, but three isolates exhibited duplications: a duplication of sp65 for DRR034478 and SRR130160, and of sp50 for DRR034476 (**Supplementary Table6**). The phylogeny that can be derived from reconstruction of the CRISPR-Cas structure confirmed the SNPs results: it reveals sporadic deletions of *cas* genes, Rv2807c, Rv2808c and Rv2813c in some isolates (**Supplementary Table7**).

IS*6110* remains a very important micro-evolutionary marker particularly in L2 that accumulated so many copies [48]. AAnc 5 strains were harboring between 14 and 22 IS*6110* copies, and two specific copies were found in almost all these L2 isolates and not in other L2 sublineages: one copy was found around position 1724419 in Rv1527c (found in 15 of these isolates) and the second one was found, around position 2041756 (found in 19 of these isolates) (**Supplementary Table5**). DRR034455, DRR034471 and DRR034476 were showing the same CRISPR structure, however harbored different missing genes (see next paragraph). Using *TB-Annotator*, 14 of the AAnc5 isolates were predicted to be drug-sensitive and four were harboring mono-resistant mutations, two were MDRs (**Supplementary Table 8**).

#### 2.3 Missing genes

Six isolates among the 20 Aanc5, apart from showing classical deletions (RD105, RD207, RD181 and PhiRv1), were harboring specific missing genes: as an example, DRR034363 had Rv1081 to 1084c deleted, DRR034416 was missing Rv1523 to Rv1526c. (**Supplementary Table7**). These deletions confirm that phylogenetically linked MTB genomes sometimes harbor strain-specific dependent deletions regions, due to recombination events and in relation to the high number of IS*6110* copies.

#### 2.4 *In Silico* VNTR copy number computation using CAST and comparison with other isolates from previous studies

Since *TB-Annotator* does not yet produce *in Silico* VNTR, we used the CAST pipeline to compute the VNTR on the 20 AAnc5 clinical isolates [55]. Apart from getting *in Silico* VNTR results, our first aim was also to compare VNTR with a very large set of isolates representative of Japan L2 biodiversity that had been published earlier [44]. Results were always partial, and no specific 15-9 VNTR signature could be obtained for the AAnc5 isolates. ETRC, QUB26 and QUB4156 could never be predicted. Depending on SRA quality, between 6 to 20 VNTR could be predicted (**Supplementary Table 9**). The VNTR results showed slight variation between isolates; eleven VNTR Loci were invariant in this collection (MIRU04, MIRU10, MIRU16, MIRU20, Mtub29, Mtub30, ETRB, MIRU24, MIRU27, Mtub34, MIRU39) whereas nine loci showed variation (MIRU02, MIRU40, Mtub21, QuB11b, ETRA, MIRU23, MIRU26, MIRU31, Mtub39). When comparing with an in-depth VNTR study performed earlier, AAnc5 was shown to belong to M10 and M37 respectively found in Russia and Singapore [51]. When comparing with a set of 5 reference japanese isolates (A05N056, ID381, 4558, 4994, 4991/M) that were described to represent the main L2 sublineages found in Japan, ID381 was sharing the same copy number with AAnc5 on 9 loci (**Supplementary Table 9**) [39, 44]. However, when comparing *in Silico* VNTR results with previous VNTR results from published studies in Korea, on the “K” strain that had been found in Korea we retrieved relatively poor similarity [38] (**Supplementary Table 9**).

#### 2.5 Comparison between *TB-Annotator* and CAST Server results on prospective Drug Susceptibility Testing and on spoligotyping results

When comparing the results obtained using *TB-Annotator* and CAST, the predictive DST results were identical (**Supplementary Table8 and Supplementary Table9**). Identical results were also obtained on the classical 43 spacers-format spoligotype reconstruction with a minor and yet unexplained discrepancy on a single spacer of a single isolate, DRR034366, for which CAST predicted SIT250 whereas TB-Annotator predicted SIT290 (**Supplementary Table9**).

#### 2.6 AAnc 5 is part of the G3 endemic L2 Ancestral strains cluster in Japan

We compared the 42 SNPs table found in the *Asian ancestral 5* sublineage (**Supplementary Table4**) with the ones found in sequences of 5 japanese reference isolates, compared to the K1-K2 epidemic strain [39]. According to the definition made by these authors 5 L2 subgroups (G1/2, G3, G4, G5/6 and M) could be defined in Japan based on 10 phylogenetically selected SNPs. From the ID381 strain, a member of the “G3” genetic group, described for the first time in Kobe and Osaka in 2006 [39], and by comparing the SNPs specific of each sublineage (Supplementary file of [39]), we came to the conclusion that the G3 group was new and did not fit with the Shitikov *et al*. classification scheme. AAnc 1, AAnc2 and AAnc3 were found to respectively match with G1/2, G4 and G5/6 in [39], however no equivalent was found for “G3”.

By comparing SNPs list, we also found that the Tochigi province AAnc5 cluster of strains was sharing 36 out of 42 SNPs with the G3:ID381 reference strain found in Kobe and Osaka (**Supplementary Table4**). 6 SNPs only (C587945T, G765140C, G1202113A, AGGGAG1476812A, G3148446C and G3820365A) were not found in the ID381 strain. We concluded that the Tochigi strains were highly likely to be historically related to the Kobe and Osaka 2006 G3 group and to the sequenced reference isolate ID381. Accordingly, we propose to retain the common SNPs described by Wada *et al*. and this study as characteristic of this sublineage, that we suggest to name *Asia Ancestral 5* (AAnc5) to fit with Shitikov *et al*. nomenclature [31]. By digging more in-depth into the comparative SNPs list between our study and the former Kobe-Osaka study, we found that DRR034489 was the closest isolate to the G3 ID381 reference sharing 40 more SNP exclusive to the G3 group, whereas another cluster of 3 genomes were more distant but were sharing 15 more SNPs with ID381 (results not shown). As mentioned above, two very distant genomes, DRR034381 and DRR130203 (417 SNPs pairwise distance) where also sharing 46 supplementary SNPs that were not found elsewhere (**Supplementary Table 5)**.

## Discussion

We describe in this paper an historical sublineage of L2 based on samples collected in central Japan, Tochigi Prefecture, former Shimotsuke province, that we coined as AAnc5. The identified sublineage is highly related to the Japanese G3 group defined in 2012 [39]. This strengthens the relevance of this sublineage and shows that it was transmitted historically in several Japanese cities. The chronology of emergence of this sublineage relatively to other L2 sublineages was positioned in Shitikov’s L2 diversification scheme. Its position relative to previously described lineages clearly showed that it should be qualified as ancestral according to the current use of this terminology and appeared just after AAnc1.

Retracing the early history of L2 in China, and South-East and in East Asia, in relation to China’s TB history, might be quite difficult [8]. Early TB outbreak history could be related to migrations of people from the 5^th^ century BC to the third century AD [84, 85]. Tuberculosis was known to be present in premodern time in Japan under the name of *rôga*, that was used in chinese medicine [86]. There are many traces in Chinese medical history texts of a disease that can be identified as tuberculosis [87]. The China-south Japan relation is known since very early in human history (1 century AD) through the *Kan-no-Wa-no-Na-no-koku*ō*in*, the seal of the King of Na, close to today’s Fukuoka; The King of Na was a vassal of the Han Emperor [88]. The hundred of small states that were present in Japan during these early years left the place to a progressive domination of the Yamato province during the Kofun period. In parallel, a south to north progression of rulers was seen in relation to the adoption of an organizational system based on the one of the Chinese empire [54]. However simultaneously, the Yamato kings were also in contact with the former Korean Baekje kingdom [89]. The Shimotsuke province, where the isolates we are describing are coming from, was in permanent contact with the Yamato court since the Kofun period [90]. This organization contributed to the creation of permanent capital cities in Japan instead of moving capitals, and a progressive development of the plains north of Edo (Tokyo), with a direct link to the former Shimotsuke known today as Tochigi province [54].

Our results on the historical connection between two locally endemic clones could only be obtained thanks to the availability of public Short Read Archives and to the development of a new bioinformatical pipeline that analyses more than 50,000 characters that builds highly resolutive phylogenetic trees, and includes repeated sequences information (PE-PPE genes, CRISPR and Insertions sequences). With the increasing power of computational analysis of genomes and using supercomputing centers, it now becomes possible to analyze not only Single-Nucleotide Polymorphisms, but also repetitive sequences, which paradoxically were at the start of molecular epidemiology [91-93], however whose analysis was almost totally supplanted by WGS [56, 57, 94]. Going more *in-depth* into the complex historical phylodynamics history of all MTC lineages (made up of various waves of expansion and extinction history) will be facilitated by the use of all available markers and the increase into study size, towards 100,000 genomes analysis, to better disentangle all the threads between ancient and recent historical event that shaped today’s pandemia, and to obtain an improved understanding of the historical pandemic in relation to ancient/modern population migrations [8, 95, 96].

In this study, we also wanted to compare the *in Silico* VNTR results of the AAnc 5 with a representative set of similar results from Kobe and Osaka [44]; however, since *in Silico* VNTR had produced sub-optimal results and since initial comparison between *in Silico* and *in Vitro* VNTR results provided limited results and is conceptually difficult using Illumina Short-reads sequencing, we did not deepen these results here since too many missing data would have jeopardize the results. The few results we obtained do not provide any information on a possible route of introduction of AAnc5 into Japan since M10 is found in Russia, whereas M37 is found in Singapore. These limited results also shows the current technical limits of *in Silico* VNTR results production; nanopore sequencing could solve this issue in a near future [97]. However when looking at SNPs dataset and the correlation between SNPs and VNTR results, it became evident that G3 and AAnc5 were sharing common ancestors.

According to its branching point, the AAnc5 lineage has emerged after AAnc1 but before AAnc 2 and 3. The very existence of a more important number of L2 ancestral clones had been guessed by the discovery of the AAnc4 [32]. The AAnc5 isolates harbored a set of unique characteristics and were shown to be distant one from the other by between 200 to 400 SNPs thus definitively excluding a contemporary outbreak history of these isolates.

Among this set of unique characteristics was the presence of a non-synonymous mutation in *rpoC*, of variations in spoligotypes as well as in VNTR structures, reinforcing progressively the evidence of historical links within the G3 and AAnc5 clusters. It is well known that *rpoC* mutations are compensatory mutations mainly found in epidemiologically-successful isolates, most of the time modern Beijing strains, that contain specific *rpoB* mutations [10, 98,Gygli, 2021 #12772]. The *rpoC* compensatory mutation could be an adaptative trait, that was suggested to explain the epidemiological success of MDR-TB L2 isolates in outbreaks such as the Central Asian or the B0/W148 outbreak in Russia [6, 99] or the epidemiological success in Georgian prisoners [36]. Merker *et al*. have shown recently in the central Asian clade that *rpoC* mutations may arise within the entire gene and authors have shown that these SNPs are found in epistasia with *rpoB* mutations, hence these mutations are designated as adaptative or compensatory [6, 10, 98]. However this is not the case here, since AAnc5 were all, except for the most recent last six isolates, drug-susceptible and hence, this *rpoC* mutation should not be considered here as a compensatory mutation, but rather as a phylogenetical marker of AAnc5 [36]. It is noteworthy that this SNP was not present in the G3 group from Kobe and Osaka. Hence, given this slight *rpoC* difference between G3 and AAnc5, it could be interesting in the future to try to search if there is a significant statistical difference between drug-resistance emergence in one or the other cluster group. We do not know if such mutations could also provide another selective advantage, independently of rifampin drug-resistance, for example on fitness, however the recent paper in prisoners in Georgia showed that compensatory mutations and patient incarceration were two independent factors associated with increased transmission, thus creating a “perfect storm” for MDR-TB transmission [36]. The precise physiological or fitness consequences of potential *rpoC* mutations in drug-sensitive isolates should be the focus of other in-depth studies; the *rpoC* SNP found in AAnc5 is non-synonymous but we did not investigate in this study if it produces a change in protein structure and has functional consequences [10]. Variations in genetic backgrounds inside L2 (intra-lineage diversity) could be linked to the early evolutionary history of AAnc5, in relation or not, to yet unknown mutator effects specific to this L2 sublineage [9].

Dating of strains diversification is important to identify the conditions that fostered past epidemics. Here, according to consensus molecular clock of TB on the middle term, the time frame of coalescence of AAnc5 sublineage landmarks could be between 280-310 years ago before present for the closest isolates, and 760-800 years ago for the farthest ones [73]. However we should be cautious in our dating estimation. Indeed, we based this estimate on L4 mutations rates [96]; we know that calibration of the molecular clock varies a lot according to samples, time-frames and lineages (between 0.04 to 2.2 SNPs per-genome-per-year) [100]. Dating can be comforted if adequate correlation exists between historical, genomic, epidemiological and demographic facts. In a former similar study, three scenarios have been tested for the dating of a L4.2 sublineage occurring in Japan and Turkey using historical, anthropological, human genetics, paleopathological events and genomics [73]. Here, for the time-being, we have few representatives of AAnc 5 within all Japan, with no independent data such as genetic backgrounds of the host, or paleontological data. We may only try to correlate our dating with historical epidemiology.

TB Transmission could have been low in early Japan during the Kofun period or later, until Japan reached a certain demography. During the 19^th^ century, the transmissible nature of TB was not admitted, it was rather believed that the disease was hereditary [101]. Within the post-Meiji society however, interest in workers’ health emerged, more for strategical than for humanitarian reasons [101]. Apart from the lack of bacteriological knowledge, fights between western and Chinese medicine lasted during centuries in Japan and bacteriology was only slowly introduced [101]. Statistics on death causes had started to be collected by the Imperial state in 1880, however tuberculosis deaths were counted in the *respiratory diseases* category, and it is only in 1883 that “pulmonary consumption” was specifically recorded as a cause of death [101, 102]. A few medical doctors at the japanese central hygiene office in 1903 were pinpointing the fact that infectious diseases first start within workers and then spread towards the general population [86]. For tuberculosis, it was linked to women working in silk and cotton spinning factories, the major driver of Japanese economy at that time [86, 103]. Hence, governments started to take TB seriously when bad health of women working in textile industry was statistically significant [86]. Mortality rate were as high as 2300 per 100,000 inhabitants in these working women, to be compared with 700 per 1000,000 in young women of the same age in the general population [101]. For a province such as Tochigi, (former Shimotsuke), with a population varying between 400,000 and 500,000 between 1720-1850 and such an incidence, around 4600-5750 cases/year could be expected in women. Then, three TB epidemic peaks are described in Japan: a first one from 1880 to 1900, a second from 1900 to 1919 with an historical peak in 1918, a third one corresponding to stabilization at a high level between 1919 till 1950 and lastly a fast decline since 1950 [101]. Women working in textile industry were paying the biggest death toll as mentioned, but men working in mining were also hit, however death causes were not distinguished from silicosis [86]. Indeed, after silk industry developed in Japan (60% of the raw silk in the world was made in Japan at the beginning of the 20th century [104], mining industry became the cornerstone of the imperial state expansion, and hence, health of male workers also became of strategic national interest for the growing military empire [86]. In the Tochigi province, the copper Ashio mine, whose exploitation started beginning of the XVII^th^ century was responsible of one-fourth of Japanese copper production in 1884; it closed down only in 1973. One of our estimation of the expansion date of the AAnc 5 is compatible wih the opening of the mine in Ashio [105]. Ashio mine could thus have been the very location of AAnc5 expansion and diversification. Looking at the historical spreading dynamics of the AAnc5 group in Japan could be done in the future by looking at finer geographical gradients of all L2 ancestral sublineages found in Japan.

Another origin of Japanese AAnc5 could be the importation of L2-ancestral clones by forced workers from Korea or China who were made to work in the mining industry from 1939 till 1945 [86]. Indeed 300,000 Koreans and 38,935 Chinese workers, mostly men were forced to work for Japan during these years [86]. We could not investigate in this study the potential distribution of G3 isolates within Japan, however our formal proof that the G3 and AAnc5 are linked and deeply rooted in Japan and that, are likely to have created independent local outbreaks in the past, is a first insight into this complex history. Future study could try to dissecate further, using combination on recent statistics on the total of epidemic cases, in combination to genomic characterization and geographical distribution, the global and the local history of ancestral L2 lineages in Japan [106].

The existence of a relatively high diversity specific to Tochigi and likely Kobe and Osaka, reminds of historical transmission restricted to a circumscribed area. Of course, such a picture of an endemic past outbreak is more easily observed in islands settings as was shown recently in New Zealand in Maori people who are hosting a specific “CS” (Colonial S-type) L4.4 Sublineage [96]. Islands are indeed excellent settings to distinguish endemic from imported species and tuberculosis history, as all infectious diseases, is definitively linked to human migration and local and global demographic history [107]. Since Japan is now a country with a very low tuberculosis prevalence rate (13 per 100,000 in 2017), it is an excellent setting to try to identify the historical events linked to past tuberculosis outbreaks [73].

## Conclusions

Thanks to *TB-Annotator*, a new bioinformatical pipeline that analyzes large amounts of the information contained in Sequence Read Archives, we mapped a local ancestral L2 sublineage onto the global MTC phylogeny, and we designated it as Asia Ancestral 5 (AAnc5), close to the formerly described G3 group in Kobe and Osaka. This sublineage now appears together with another one in Shitikov’s evolutionary scheme. This sublineage possesses many specific characters allowing to distinguish it from all other historical clones described so far for the “Beijing” (Lineage 2). This finding opens new ways of research, to look for the history of spreading of this cluster within all Japan, and further in relation to Korea, to South China or elsewhere in South-East Asia and East Asia. This pipeline will permit new investigations on the spatial and dynamics history of tuberculosis to be made.

## Data Availability

All data are publicly available or contained in the article as supplementary files.

## Legend to Supplementary Material

**Supplementary Table1**: List of all 190 SRAs of the L2 lineage investigated in this study, with a majority of L2 from central Tochigi, and other L2 representing the diversity of L2 sublineages described so far, with the exception of Pacific RD150, main SNPs assessed, and assignment to Shitikov classification scheme or newly assigned for Asia Ancestral 4 and 5.

**Supplementary Table2**: List of SNPs catalogs and/or other markers, used to classify SRAs in *TB-Annotator*

**Supplementary Table3**: Full Excel Matrix results produced on the L2 Lineage for the 190 L2 isolates and list of 680 SRAs used in the study (Sheet 1: General)

**Supplementary Table4**: list of 42 SNPs specific of Asia Ancestral 5

**Supplementary Table5**: SNPs Pairwise distance between the 20 Asia Ancestral 5 isolates, extra list of 46 SNPs found between two AAnc5 isolates.

**Supplementary Table6**: IS*6110* insertion sites on the 20 Asia Ancestral 5 isolates

**Supplementary Table7**: summary of missing genes for the 20 Asia Ancestral 5 isolates

**Supplementary Table8**: SNPs detected in drug resistance genes in the 20 Asia Ancestral 5 isolates

**Supplementary Table9**: *in Silico* VNTR results on the 20 Asia Ancestral 5 and comparison of predictive DST and spoligotyping using CAST or *TB-Annotator*

## References

1. WHO. Global Tuberculosis Report 2020. WHO, Geneva, Switzerland, 2020 2020, Oct. 15th. Report No.

2. Devaux I, Kremer K, Heersma H, Van Soolingen D. Clusters of multidrug-resistant Mycobacterium tuberculosis cases, Europe. Emerg Infect Dis. 2009;15(7):1052–60. Epub 2009/07/25. doi: 10.3201/eid1507.080994. PubMed PMID: 19624920; PubMed Central PMCID: PMCPMC2744220.

3. Affolabi D, Faihun F, Sanoussi N, Anyo G, Shamputa IC, Rigouts L, et al. Possible outbreak of streptomycin-resistant Mycobacterium tuberculosis Beijing in Benin. Emerg Infect Dis. 2009;15(7):1123–5. Epub 2009/07/25. doi: 10.3201/eid1507.080697. PubMed PMID: 19624936; PubMed Central PMCID: PMCPMC2744252.

4. Casali N, Nikolayevskyy V, Balabanova Y, Harris SR, Ignatyeva O, Kontsevaya I, et al. Evolution and transmission of drug-resistant tuberculosis in a Russian population. Nature genetics. 2014;46(3):279–86. Epub 2014/01/28. doi: 10.1038/ng.2878. PubMed PMID: 24464101; PubMed Central PMCID: PMC3939361.

5. Iwamoto T, Grandjean L, Arikawa K, Nakanishi N, Caviedes L, Coronel J, et al. Genetic diversity and transmission characteristics of Beijing family strains of Mycobacterium tuberculosis in Peru. PLoS One. 2012;7(11):e49651. Epub 2012/11/28. doi: 10.1371/journal.pone.0049651. PubMed PMID: 23185395; PubMed Central PMCID: PMCPMC3504116.

6. Merker M, Barbier M, Cox H, Rasigade JP, Feuerriegel S, Kohl TA, et al. Compensatory evolution drives multidrug-resistant tuberculosis in Central Asia. eLife. 2018;7. Epub 2018/10/31. doi: 10.7554/eLife.38200. PubMed PMID: 30373719; PubMed Central PMCID: PMCPMC6207422.

7. Liu Q, Luo T, Dong X, Sun G, Liu Z, Gan M, et al. Genetic features of Mycobacterium tuberculosis modern Beijing sublineage. Emerg Microbes Infect. 2016;5:e14. Epub 2016/02/26. doi: 10.1038/emi.2016.14. PubMed PMID: 26905026; PubMed Central PMCID: PMCPMC4777927.

8. Liu Q, Ma A, Wei L, Pang Y, Wu B, Luo T, et al. China’s tuberculosis epidemic stems from historical expansion of four strains of Mycobacterium tuberculosis. Nat Ecol Evol. 2018;2(12):1982–92. Epub 2018/11/07. doi: 10.1038/s41559-018-0680-6. PubMed PMID: 30397300; PubMed Central PMCID: PMCPMC6295914.

9. Rad ME, Bifani P, Martin C, Kremer K, Samper S, Rauzier J, et al. Mutations in putative mutator genes of Mycobacterium tuberculosis strains of the W-Beijing family. Emerg Infect Dis. 2003;9(7):838–45.

10. Comas I, Borrell S, Roetzer A, Rose G, Malla B, Kato-Maeda M, et al. Whole-genome sequencing of rifampicin-resistant Mycobacterium tuberculosis strains identifies compensatory mutations in RNA polymerase genes. Nature genetics. 2011;44(1):106–10. Epub 2011/12/20. doi: 10.1038/ng.1038. PubMed PMID: 22179134; PubMed Central PMCID: PMCPMC3246538.

11. Klotoe BJ, Kacimi S, Costa-Conceicao E, Gomes HM, Barcellos RB, Panaiotov S, et al. Genomic characterization of MDR/XDR-TB in Kazakhstan by a combination of high-throughput methods predominantly shows the ongoing transmission of L2/Beijing 94-32 central Asian/Russian clusters. BMC Infect Dis. 2019;19(1):553. doi: 10.1186/s12879-019-4201-2. PubMed PMID: 31234780.

12. Droznin M, Johnson A, Johnson AM. Multidrug resistant tuberculosis in prisons located in former Soviet countries: A systematic review. PLoS One. 2017;12(3):e0174373. Epub 2017/03/24. doi: 10.1371/journal.pone.0174373. PubMed PMID: 28334036; PubMed Central PMCID: PMCPMC5363920.

13. Vidya C, Prabheesh K. Implications of COVID-19 Pandemic on the Global Trade Networks. EMERGING MARKETS FINANCE AND TRADE. 2020;2020(56):2408–21.

14. Glaziou P. Predicted impact of the COVID-19 pandemic on global tuberculosis deaths in 2020. medRxiv. 2020.

15. Nicita A, Razo C. China: The rise of a trade titan: UNCTAD; 2021 [cited 2021 May 31st, 2021]. Available from: https://unctad.org/news/china-rise-trade-titan.

16. van Soolingen D, Qian L, de Haas PE, Douglas JT, Traore H, Portaels F, et al. Predominance of a single genotype of Mycobacterium tuberculosis in countries of east Asia. J Clin Microbiol. 1995;33(12):3234-8. PubMed PMID: 8586708.

17. Plikaytis BB, Musser JM, Bifani P, Kreiswirth BN, Emben JDAv, Crawford JT, et al., editors. New York city Mycobacterium tuberculosis W family shares common genetic markers with the Mycobacterium tuberculosis Beijing family. 17th Annual Meeting of the European Society of Mycobacteriology; 1996; Paris.

18. Plikaytis BB, Marden JL, Crawford JT, Woodley CL, Butler WR, Shinnick TM. Multiplex PCR assay specific for the multidrug-resistant strain W of Mycobacterium tuberculosis. J Clin Microbiol. 1994;32(6):1542-6. PubMed PMID: 7915723; PubMed Central PMCID: PMC264034.

19. Kurepina NE, Sreevatsan S, Plikaytis BB, Bifani PB, Connell ND, Donneelly RJ, et al. Characterization of the phylogenetic distribution and chromosomal insertion sites of five IS6110 elements in Mycobacterium tuberculosis: non random integration in the dnaA-dnaN region. Tubercle Lung Dis. 1998;79:31–42.

20. Bifani PJ, Mathema B, Kurepina NE, Kreiswirth BN. Global dissemination of the Mycobacterium tuberculosis W-Beijing family strains. Trends Microbiol. 2002;10(1):45-52. PubMed PMID: 11755085.

21. Mokrousov I, Ly HM, Otten T, Lan NN, Vyshnevskyi B, Hoffner S, et al. Origin and primary dispersal of the Mycobacterium tuberculosis Beijing genotype: clues from human phylogeography. Genome Res. 2005;15(10):1357–64. Epub 2005/09/20. doi: 10.1101/gr.3840605. PubMed PMID: 16169923; PubMed Central PMCID: PMCPMC1240077.

22. Tsolaki AG, Gagneux S, Pym AS, Goguet de la Salmoniere YO, Kreiswirth BN, Van Soolingen D, et al. Genomic deletions classify the Beijing/W strains as a distinct genetic lineage of Mycobacterium tuberculosis. J Clin Microbiol. 2005;43(7):3185–91. Epub 2005/07/08. doi: 10.1128/JCM.43.7.3185-3191.2005. PubMed PMID: 16000433; PubMed Central PMCID: PMCPMC1169157.

23. Merker M, Blin C, Mona S, Duforet-Frebourg N, Lecher S, Willery E, et al. Evolutionary history and global spread of the Mycobacterium tuberculosis Beijing lineage. Nature genetics. 2015;47(3):242–9. doi: 10.1038/ng.3195. PubMed PMID: 25599400.

24. Mokrousov I, Narvskaya O, Vyazovaya A, Otten T, Jiao WW, Gomes LL, et al. Russian “successful” clone B0/W148 of Mycobacterium tuberculosis Beijing genotype: a multiplex PCR assay for rapid detection and global screening. J Clin Microbiol. 2012;50(11):3757–9. Epub 2012/08/31. doi: 10.1128/JCM.02001-12. PubMed PMID: 22933595; PubMed Central PMCID: PMCPMC3486266.

25. Mokrousov I, Narvskaya O, Otten T, Vyazovaya A, Limeschenko E, Steklova L, et al. Phylogenetic reconstruction within Mycobacterium tuberculosis Beijing genotype in northwestern Russia. Res Microbiol. 2002;153(10):629–37.

26. Kremer K, Glynn JR, Lillebaek T, Niemann S, Kurepina NE, Kreiswirth BN, et al. Definition of the Beijing/W lineage of Mycobacterium tuberculosis on the basis of genetic markers. J Clin Microbiol. 2004;42(9):4040–9.

27. Luo T, Comas I, Luo D, Lu B, Wu J, Wei L, et al. Southern East Asian origin and coexpansion of Mycobacterium tuberculosis Beijing family with Han Chinese. Proceedings of the National Academy of Sciences of the United States of America. 2015;112(26):8136–41. doi: 10.1073/pnas.1424063112. PubMed PMID: 26080405; PubMed Central PMCID: PMCPMC4491734.

28. Srilohasin P, Prammananan T, Faksri K, Phelan JE, Suriyaphol P, Kamolwat P, et al. Genomic evidence supporting the clonal expansion of extensively drug-resistant tuberculosis bacteria belonging to a rare proto-Beijing genotype. Emerg Microbes Infect. 2020;9(1):2632–41. Epub 2020/11/19. doi: 10.1080/22221751.2020.1852891. PubMed PMID: 33205698; PubMed Central PMCID: PMCPMC7738298.

29. Coll F, McNerney R, Guerra-Assuncao JA, Glynn JR, Perdigao J, Viveiros M, et al. A robust SNP barcode for typing Mycobacterium tuberculosis complex strains. Nature communications. 2014;5:4812. doi: 10.1038/ncomms5812. PubMed PMID: 25176035; PubMed Central PMCID: PMC4166679.

30. Yin QQ, Liu HC, Jiao WW, Li QJ, Han R, Tian JL, et al. Evolutionary History and Ongoing Transmission of Phylogenetic Sublineages of Mycobacterium tuberculosis Beijing Genotype in China. Scientific reports. 2016;6:34353. Epub 2016/09/30. doi: 10.1038/srep34353. PubMed PMID: 27681182; PubMed Central PMCID: PMCPmc5041183.

31. Shitikov E, Kolchenko S, Mokrousov I, Bespyatykh J, Ischenko D, Ilina E, et al. Evolutionary pathway analysis and unified classification of East Asian lineage of Mycobacterium tuberculosis. Scientific reports. 2017;7(1):9227. doi: 10.1038/s41598-017-10018-5. PubMed PMID: 28835627; PubMed Central PMCID: PMCPMC5569047.

32. Ajawatanawong P, Yanai H, Smittipat N, Disratthakit A, Yamada N, Miyahara R, et al. A novel Ancestral Beijing sublineage of Mycobacterium tuberculosis suggests the transition site to Modern Beijing sublineages. Scientific reports. 2019;9(1):13718. Epub 2019/09/25. doi: 10.1038/s41598-019-50078-3. PubMed PMID: 31548561; PubMed Central PMCID: PMCPMC6757101.

33. Shitikov EA, Bespyatykh JA, Ischenko DS, Alexeev DG, Karpova IY, Kostryukova ES, et al. Unusual large-scale chromosomal rearrangements in Mycobacterium tuberculosis Beijing B0/W148 cluster isolates. PLoS One. 2014;9(1):e84971. doi: 10.1371/journal.pone.0084971. PubMed PMID: 24416324; PubMed Central PMCID: PMCPMC3885621.

34. Wang WF, Lu MJ, Cheng TR, Tang YC, Teng YC, Hwa TY, et al. Genomic Analysis of Mycobacterium tuberculosis Isolates and Construction of a Beijing Lineage Reference Genome. Genome biology and evolution. 2020;12(2):3890–905. Epub 2020/01/24. doi: 10.1093/gbe/evaa009. PubMed PMID: 31971587.

35. Karmakar M, Trauer JM, Ascher DB, Denholm JT. Hyper transmission of Beijing lineage Mycobacterium tuberculosis: Systematic review and meta-analysis. The Journal of infection. 2019;79(6):572–81. Epub 2019/10/05. doi: 10.1016/j.jinf.2019.09.016. PubMed PMID: 31585190.

36. Gygli SM, Loiseau C, Jugheli L, Adamia N, Trauner A, Reinhard M, et al. Prisons as ecological drivers of fitness-compensated multidrug-resistant Mycobacterium tuberculosis. Nat Med. 2021. Epub 2021/05/26. doi: 10.1038/s41591-021-01358-x. PubMed PMID: 34031604.

37. Mestre O, Luo T, Dos Vultos T, Kremer K, Murray A, Namouchi A, et al. Phylogeny of Mycobacterium tuberculosis Beijing strains constructed from polymorphisms in genes involved in DNA replication, recombination and repair. PLoS One. 2011;6(1):e16020. PubMed PMID: 21283803.

38. Kang HY, Wada T, Iwamoto T, Maeda S, Murase Y, Kato S, et al. Phylogeographical particularity of the Mycobacterium tuberculosis Beijing family in South Korea based on international comparison with surrounding countries. J Med Microbiol. 2010;59(Pt 10):1191–7. Epub 2010/06/26. doi: 10.1099/jmm.0.022103-0. PubMed PMID: 20576748.

39. Wada T, Iwamoto T, Hase A, Maeda S. Scanning of genetic diversity of evolutionarily sequential Mycobacterium tuberculosis Beijing family strains based on genome wide analysis. Infect Genet Evol. 2012;12(7):1392–6. Epub 2012/05/15. doi: 10.1016/j.meegid.2012.04.029. PubMed PMID: 22580240.

40. Park YK, Kang H, Yoo H, Lee SH, Roh H, Kim HJ, et al. Whole-Genome Sequence of Mycobacterium tuberculosis Korean Strain KIT87190. Genome announcements. 2014;2(5). Epub 2014/11/02. doi: 10.1128/genomeA.01103-14. PubMed PMID: 25359915; PubMed Central PMCID: PMCPMC4214991.

41. Wada T, Iwamoto T, Tamaru A, Seto J, Ahiko T, Yamamoto K, et al. Clonality and micro-diversity of a nationwide spreading genotype of Mycobacterium tuberculosis in Japan. PLoS One. 2015;10(3):e0118495. Epub 2015/03/04. doi: 10.1371/journal.pone.0118495. PubMed PMID: 25734518; PubMed Central PMCID: PMCPMC4348518.

42. Mizukoshi F, Miyoshi-Akiyama T, Iwai H, Suzuki T, Kiritani R, Kirikae T, et al. Genetic diversity of Mycobacterium tuberculosis isolates from Tochigi prefecture, a local region of Japan. BMC Infect Dis. 2017;17(1):365. Epub 2017/05/27. doi: 10.1186/s12879-017-2457-y. PubMed PMID: 28545488; PubMed Central PMCID: PMCPMC5445273.

43. Mokrousov I, Narvskaya O, Vyazovaya A, Millet J, Otten T, Vishnevsky B, et al. Mycobacterium tuberculosis Beijing genotype in Russia: in search of informative variable-number tandem-repeat loci. J Clin Microbiol. 2008;46(11):3576–84. Epub 2008/08/30. doi: 10.1128/JCM.00414-08. PubMed PMID: 18753356; PubMed Central PMCID: PMCPMC2576596.

44. Iwamoto T, Fujiyama R, Yoshida S, Wada T, Shirai C, Kawakami Y. Population structure dynamics of Mycobacterium tuberculosis Beijing strains during past decades in Japan. J Clin Microbiol. 2009;47(10):3340-3. PubMed PMID: 19710282.

45. Allix-Beguec C, Wahl C, Hanekom M, Nikolayevskyy V, Drobniewski F, Maeda S, et al. Proposal of a consensus set of hypervariable mycobacterial interspersed repetitive-unit-variable-number tandem-repeat loci for subtyping of Mycobacterium tuberculosis Beijing isolates. J Clin Microbiol. 2014;52(1):164–72. Epub 2013/11/01. doi: 10.1128/JCM.02519-13. PubMed PMID: 24172154; PubMed Central PMCID: PMCPMC3911419.

46. Casali N, Nikolayevskyy V, Balabanova Y, Ignatyeva O, Kontsevaya I, Harris SR, et al. Microevolution of extensively drug-resistant tuberculosis in Russia. Genome Res. 2012;22(4):735–45. Epub 2012/02/02. doi: 10.1101/gr.128678.111. PubMed PMID: 22294518; PubMed Central PMCID: PMC3317155.

47. Roychowdhury T, Mandal S, Bhattacharya A. Analysis of IS6110 insertion sites provide a glimpse into genome evolution of Mycobacterium tuberculosis. Scientific reports. 2015;5:12567. doi: 10.1038/srep12567. PubMed PMID: 26215170; PubMed Central PMCID: PMC4517164.

48. Shitikov E, Guliaev A, Bespyatykh J, Malakhova M, Kolchenko S, Smirnov G, et al. The role of IS6110 in micro-and macroevolution of Mycobacterium tuberculosis lineage 2. Molecular phylogenetics and evolution. 2019;139:106559. Epub 2019/07/16. doi: 10.1016/j.ympev.2019.106559. PubMed PMID: 31302224.

49. Ford CB, Shah RR, Maeda MK, Gagneux S, Murray MB, Cohen T, et al. Mycobacterium tuberculosis mutation rate estimates from different lineages predict substantial differences in the emergence of drug-resistant tuberculosis. Nature genetics. 2013;45(7):784–90. Epub 2013/06/12. doi: 10.1038/ng.2656. PubMed PMID: 23749189; PubMed Central PMCID: PMCPMC3777616.

50. Hirsh AE, Tsolaki AG, DeRiemer K, Feldman MW, Small PM. Stable association between strains of Mycobacterium tuberculosis and their human host populations. Proceedings of the National Academy of Sciences of the United States of America. 2004;101(14):4871–6. Epub 2004/03/26. doi: 10.1073/pnas.0305627101. PubMed PMID: 15041743; PubMed Central PMCID: PMCPMC387341.

51. Mokrousov I. Genetic geography of Mycobacterium tuberculosis Beijing genotype: a multifacet mirror of human history? Infect Genet Evol. 2008;8(6):777–85. doi: 10.1016/j.meegid.2008.07.003. PubMed PMID: 18691674.

52. Abdulla MA, Ahmed I, Assawamakin A, Bhak J, Brahmachari SK, Calacal GC, et al. Mapping human genetic diversity in Asia. Science. 2009;326(5959):1541–5. Epub 2009/12/17. doi: 10.1126/science.1177074. PubMed PMID: 20007900.

53. Wada T, Iwamoto T. Allelic diversity of variable number of tandem repeats provides phylogenetic clues regarding the Mycobacterium tuberculosis Beijing family. Infect Genet Evol. 2009;9(5):921–6. Epub 2009/06/23. doi: 10.1016/j.meegid.2009.06.012. PubMed PMID: 19540936.

54. Wikipedia. Japan History https://en.wikipedia.org/wiki/History_of_Japan2021 [cited 2021 May 31st 2021]. Available from: https://en.wikipedia.org/wiki/History_of_Japan.

55. Iwai H, Kato-Miyazawa M, Kirikae T, Miyoshi-Akiyama T. CASTB (the comprehensive analysis server for the Mycobacterium tuberculosis complex): A publicly accessible web server for epidemiological analyses, drug-resistance prediction and phylogenetic comparison of clinical isolates. Tuberculosis (Edinb). 2015;95(6):843–4. doi: 10.1016/j.tube.2015.09.002. PubMed PMID: 26542225.

56. Freschi L, Vargas R, Hussain A, Kamal SMM, Skrahina A, Tahseen S, et al. Population structure, biogeography and transmissibility of <em>Mycobacterium tuberculosis</em>. bioRxiv. 2020:2020.09.29.293274. doi: 10.1101/2020.09.29.293274.

57. Napier G, Campino S, Merid Y, Abebe M, Woldeamanuel Y, Aseffa A, et al. Robust barcoding and identification of Mycobacterium tuberculosis lineages for epidemiological and clinical studies. Genome medicine. 2020;12(1):114. Epub 2020/12/16. doi: 10.1186/s13073-020-00817-3. PubMed PMID: 33317631; PubMed Central PMCID: PMCPMC7734807.

58. Refrégier G, Sola C, Guyeux C. Unexpected diversity of CRISPR unveils some evolutionary patterns of repeated sequences in Mycobacterium tuberculosis. BMC genomics. 2020;21:841. doi: https://doi.org/10.1186/s12864-020-07178-6.

59. Guyeux C, Sola C, Nous C, Refregier G. CRISPRbuilder-TB: “CRISPR-builder for tuberculosis”. Exhaustive reconstruction of the CRISPR locus in mycobacterium tuberculosis complex using SRA. PLoS computational biology. 2021;17(3):e1008500. Epub 2021/03/06. doi: 10.1371/journal.pcbi.1008500. PubMed PMID: 33667225; PubMed Central PMCID: PMCPMC7968741.

60. Sreevatsan S, Pan X, Stockbauer KE, Connell ND, Kreiswirth BN, Whittam TS, et al. Restricted structural gene polymorphism in the Mycobacterium tuberculosis complex indicates evolutionarily recent global dissemination. Proceedings of the National Academy of Sciences of the United States of America. 1997;94(18):9869-74. PubMed PMID: 9275218.

61. Malm S, Linguissi LS, Tekwu EM, Vouvoungui JC, Kohl TA, Beckert P, et al. New Mycobacterium tuberculosis Complex Sublineage, Brazzaville, Congo. Emerg Infect Dis. 2017;23(3):423–9. doi: 10.3201/eid2303.160679. PubMed PMID: 28221129; PubMed Central PMCID: PMCPMC5382753.

62. Filliol I, Motiwala AS, Cavatore M, Qi W, Hazbon MH, Bobadilla del Valle M, et al. Global phylogeny of Mycobacterium tuberculosis based on single nucleotide polymorphism (SNP) analysis: insights into tuberculosis evolution, phylogenetic accuracy of other DNA fingerprinting systems, and recommendations for a minimal standard SNP set. Journal of bacteriology. 2006;188(2):759-72. PubMed PMID: 16385065.

63. Borrell S, Trauner A, Brites D, Rigouts L, Loiseau C, Coscolla M, et al. Reference set of Mycobacterium tuberculosis clinical strains: A tool for research and product development. PLoS One. 2019;14(3):e0214088. Epub 2019/03/26. doi: 10.1371/journal.pone.0214088. PubMed PMID: 30908506; PubMed Central PMCID: PMCPMC6433267.

64. Faksri K, Xia E, Tan JH, Teo YY, Ong RT. In silico region of difference (RD) analysis of Mycobacterium tuberculosis complex from sequence reads using RD-Analyzer. BMC genomics. 2016;17(1):847. Epub 2016/11/04. doi: 10.1186/s12864-016-3213-1. PubMed PMID: 27806686; PubMed Central PMCID: PMCPMC5093977.

65. Brosch R, Gordon SV, Marmiesse M, Brodin P, Buchrieser C, Eiglmeier K, et al. A new evolutionary scenario for the Mycobacterium tuberculosis complex. Proceedings of the National Academy of Sciences of the United States of America. 2002;99(6):3684–9. Epub 2002/03/14. doi: 10.1073/pnas.052548299. PubMed PMID: 11891304; PubMed Central PMCID: PMCPMC122584.

66. Palittapongarnpim P, Ajawatanawong P, Viratyosin W, Smittipat N, Disratthakit A, Mahasirimongkol S, et al. Evidence for Host-Bacterial Co-evolution via Genome Sequence Analysis of 480 Thai Mycobacterium tuberculosis Lineage 1 Isolates. Scientific reports. 2018;8(1):11597. Epub 2018/08/04. doi: 10.1038/s41598-018-29986-3. PubMed PMID: 30072734; PubMed Central PMCID: PMCPMC6072702.

67. Coscolla M, Brites D, Menardo F, Loiseau C, Borrell S, Otchere ID, et al. Phylogenomics of Mycobacterium africanum reveals a new lineage and a complex evolutionary history. bioRxiv [Internet]. 2020. Available from: https://doi.org/10.1101/2020.06.10.141788.

68. Stucki D, Brites D, Jeljeli L, Coscolla M, Liu Q, Trauner A, et al. Mycobacterium tuberculosis lineage 4 comprises globally distributed and geographically restricted sublineages. Nature genetics. 2016;48(12):1535–43. doi: 10.1038/ng.3704. PubMed PMID: 27798628.

69. Homolka S, Projahn M, Feuerriegel S, Ubben T, Diel R, Nubel U, et al. High Resolution Discrimination of Clinical Mycobacterium tuberculosis Complex Strains Based on Single Nucleotide Polymorphisms. PLoS One. 2012;7(7):e39855. Epub 2012/07/07. doi: 10.1371/journal.pone.0039855. PubMed PMID: 22768315; PubMed Central PMCID: PMC3388094.

70. Comas I, Homolka S, Niemann S, Gagneux S. Genotyping of genetically monomorphic bacteria: DNA sequencing in Mycobacterium tuberculosis highlights the limitations of current methodologies. PLoS One. 2009;4(11):e7815. Epub 2009/11/17. doi: 10.1371/journal.pone.0007815. PubMed PMID: 19915672; PubMed Central PMCID: PMCPMC2772813.

71. Comas I, Coscolla M, Luo T, Borrell S, Holt KE, Kato-Maeda M, et al. Out-of-Africa migration and Neolithic coexpansion of Mycobacterium tuberculosis with modern humans. Nature genetics. 2013;45(10):1176–82. Epub 2013/09/03. doi: 10.1038/ng.2744. PubMed PMID: 23995134.

72. Mokrousov I, Shitikov E, Skiba Y, Kolchenko S, Chernyaeva E, Vyazovaya A. Emerging peak on the phylogeographic landscape of Mycobacterium tuberculosis in West Asia: Definitely smoke, likely fire. Molecular phylogenetics and evolution. 2017;116:202–12. doi: 10.1016/j.ympev.2017.09.002. PubMed PMID: 28893611.

73. Refregier G, Abadia E, Matsumoto T, Ano H, Takashima T, Tsuyuguchi I, et al. Turkish and Japanese Mycobacterium tuberculosis sublineages share a remote common ancestor. Infect Genet Evol. 2016;45:461–73. doi: 10.1016/j.meegid.2016.10.009. PubMed PMID: 27746295.

74. Tantivitayakul P, Juthayothin T, Ruangchai W, Smittipat N, Disratthakit A, Mahasirimongkol S, et al. Identification and in silico functional prediction of lineage-specific SNPs distributed in DosR-related proteins and resuscitation-promoting factor proteins of Mycobacterium tuberculosis. Heliyon. 2020;6(12):e05744. Epub 2020/12/29. doi: 10.1016/j.heliyon.2020.e05744. PubMed PMID: 33364506; PubMed Central PMCID: PMCPMC7753917.

75. Ates LS, Dippenaar A, Sayes F, Pawlik A, Bouchier C, Ma L, et al. Unexpected Genomic and Phenotypic Diversity of Mycobacterium africanum Lineage 5 Affects Drug Resistance, Protein Secretion, and Immunogenicity. Genome biology and evolution. 2018;10(8):1858–74. Epub 2018/07/17. doi: 10.1093/gbe/evy145. PubMed PMID: 30010947; PubMed Central PMCID: PMCPMC6071665.

76. Lipworth S, Jajou R, de Neeling A, Bradley P, van der Hoek W, Maphalala G, et al. SNP-IT Tool for Identifying Subspecies and Associated Lineages of Mycobacterium tuberculosis Complex. Emerg Infect Dis. 2019;25(3):482–8. Epub 2019/02/23. doi: 10.3201/eid2503.180894. PubMed PMID: 30789126; PubMed Central PMCID: PMCPMC6390766.

77. Kohl TA, Utpatel C, Schleusener V, De Filippo MR, Beckert P, Cirillo DM, et al. MTBseq: a comprehensive pipeline for whole genome sequence analysis of Mycobacterium tuberculosis complex isolates. PeerJ. 2018;6:e5895. Epub 2018/11/28. doi: 10.7717/peerj.5895. PubMed PMID: 30479891; PubMed Central PMCID: PMCPMC6238766.

78. Coll F, Phelan J, Hill-Cawthorne GA, Nair MB, Mallard K, Ali S, et al. Genome-wide analysis of multi-and extensively drug-resistant Mycobacterium tuberculosis. Nature genetics. 2018;50(2):307–16. doi: 10.1038/s41588-017-0029-0. PubMed PMID: 29358649.

79. Coker OO, Chaiprasert A, Ngamphiw C, Tongsima S, Regmi SM, Clark TG, et al. Genetic signatures of Mycobacterium tuberculosis Nonthaburi genotype revealed by whole genome analysis of isolates from tuberculous meningitis patients in Thailand. PeerJ. 2016;4:e1905. Epub 2016/04/27. doi: 10.7717/peerj.1905. PubMed PMID: 27114869; PubMed Central PMCID: PMCPMC4841212.

80. Koster K, Largen A, Foster JT, Drees KP, Qian L, Desmond EP, et al. Whole genome SNP analysis suggests unique virulence factor differences of the Beijing and Manila families of Mycobacterium tuberculosis found in Hawaii. PLoS One. 2018;13(7):e0201146. Epub 2018/07/24. doi: 10.1371/journal.pone.0201146. PubMed PMID: 30036392; PubMed Central PMCID: PMCPMC6056056.

81. Couvin D, David A, Zozio T, Rastogi N. Macro-geographical specificities of the prevailing tuberculosis epidemic as seen through SITVIT2, an updated version of the Mycobacterium tuberculosis genotyping database. Infect Genet Evol. 2019;72:31–43. Epub 2018/12/30. doi: 10.1016/j.meegid.2018.12.030. PubMed PMID: 30593925.

82. Stamatakis A. RAxML version 8: a tool for phylogenetic analysis and post-analysis of large phylogenies. Bioinformatics. 2014;30(9):1312–3. Epub 2014/01/24. doi: 10.1093/bioinformatics/btu033. PubMed PMID: 24451623; PubMed Central PMCID: PMCPMC3998144.

83. Kloepper TH, Huson DH. Drawing explicit phylogenetic networks and their integration into SplitsTree. BMC evolutionary biology. 2008;8:22. Epub 2008/01/26. doi: 10.1186/1471-2148-8-22. PubMed PMID: 18218099; PubMed Central PMCID: PMCPMC2253509.

84. Suzuki T, Fujita H, Choi JG. Brief communication: new evidence of tuberculosis from prehistoric Korea-Population movement and early evidence of tuberculosis in far East Asia. American journal of physical anthropology. 2008;136(3):357–60. doi: 10.1002/ajpa.20811. PubMed PMID: 18322918.

85. Suzuki T, Inoue T. Earliest Evidence of Spinal Tuberculosis from the Aneolithic Yayoi Period in Japan. International Journal of Osteoarchaeology. 2007;17:392–492.

86. Thomann B. L’hygiène nationale, la société civile et la reconnaissance de la silicose comme maladie professionnelle au Japon (1868-1960). Revue d’Histoire Moderne et Contemporaine. 2009;2009/1(56-1):142–76.

87. Barberis I, Bragazzi NL, Galluzzo L, Martini M. The history of tuberculosis: from the first historical records to the isolation of Koch’s bacillus. J Prev Med Hyg. 2017;58(1):E9–E12. Epub 2017/05/19. PubMed PMID: 28515626; PubMed Central PMCID: PMCPMC5432783.

88. Wikipedia. King of Na gold Seal https://en.wikipedia.org/wiki/King_of_Na_gold_seal?oldid=495551629; 2021 [cited 2021 2021, May 31st].

89. Wikipedia. Three kingdoms of Korea https://en.wikipedia.org/wiki/Three_Kingdoms_of_Korea2021 [cited 2021 May 31st, 2021]. Available from: https://en.wikipedia.org/wiki/Three_Kingdoms_of_Korea.

90. Anonymous. Shimotsuke province Wikipedia: wikipedia; [cited 2021 2021, Jun 9th]. Available from: https://en.wikipedia.org/wiki/Shimotsuke_Province.

91. van Embden JD, Cave MD, Crawford JT, Dale JW, Eisenach KD, Gicquel B, et al. Strain identification of Mycobacterium tuberculosis by DNA fingerprinting: recommendations for a standardized methodology. J Clin Microbiol. 1993;31(2):406-9. PubMed PMID: 8381814.

92. Kamerbeek J, Schouls L, Kolk A, van Agterveld M, van Soolingen D, Kuijper S, et al. Simultaneous detection and strain differentiation of Mycobacterium tuberculosis for diagnosis and epidemiology. J Clin Microbiol. 1997;35(4):907-14. PubMed PMID: 9157152.

93. Supply P, Allix C, Lesjean S, Cardoso-Oelemann M, Rusch-Gerdes S, Willery E, et al. Proposal for standardization of optimized mycobacterial interspersed repetitive unit-variable-number tandem repeat typing of Mycobacterium tuberculosis. J Clin Microbiol. 2006;44(12):4498–510. Epub 2006/09/29. doi: 10.1128/JCM.01392-06. PubMed PMID: 17005759; PubMed Central PMCID: PMCPMC1698431.

94. Menardo F, Rutaihwa L, Zwyer M, Borrell S, Comas I, ConceiÁ„o E, et al. Local adaptation in populations of Mycobacterium tuberculosis endemic to the Indian Ocean Rim [version 2; peer review: 2 approved]. F1000Research. 2021;10(60). doi: 10.12688/f1000research.28318.1.

95. Pepperell CS, Granka JM, Alexander DC, Behr MA, Chui L, Gordon J, et al. Dispersal of Mycobacterium tuberculosis via the Canadian fur trade. Proceedings of the National Academy of Sciences of the United States of America. 2011;108(16):6526–31. Epub 2011/04/06. doi: 10.1073/pnas.1016708108. PubMed PMID: 21464295; PubMed Central PMCID: PMCPMC3080970.

96. Mulholland CV, Shockey AC, L. AH, Cursons RT, O’Toole RF, Gautam SS, et al. Dispersal of Mycobacterium tuberculosis driven by historical european trade in the South Pacific. Frontiers in microbiology. 2019;doi: 10.3389/fmicb.2019.02778. Epub 04 December 2019.

97. Bainomugisa A, Duarte T, Lavu E, Pandey S, Coulter C, Marais BJ, et al. A complete high-quality MinION nanopore assembly of an extensively drug-resistant Mycobacterium tuberculosis Beijing lineage strain identifies novel variation in repetitive PE/PPE gene regions. Microb Genom. 2018;4(7). Epub 2018/06/16. doi: 10.1099/mgen.0.000188. PubMed PMID: 29906261; PubMed Central PMCID: PMCPMC6113869.

98. de Vos M, Muller B, Borrell S, Black PA, van Helden PD, Warren RM, et al. Putative compensatory mutations in the rpoC gene of rifampin-resistant Mycobacterium tuberculosis are associated with ongoing transmission. Antimicrob Agents Chemother. 2013;57(2):827–32. Epub 2012/12/05. doi: 10.1128/AAC.01541-12. PubMed PMID: 23208709; PubMed Central PMCID: PMC3553702.

99. Li QJ, Jiao WW, Yin QQ, Xu F, Li JQ, Sun L, et al. Compensatory Mutations of Rifampin Resistance Are Associated with Transmission of Multidrug-Resistant Mycobacterium tuberculosis Beijing Genotype Strains in China. Antimicrob Agents Chemother. 2016;60(5):2807–12. Epub 2016/02/24. doi: 10.1128/aac.02358-15. PubMed PMID: 26902762; PubMed Central PMCID: PMCPmc4862491.

100. Menardo F, Duchene S, Brites D, Gagneux S. The molecular clock of Mycobacterium tuberculosis. PLoS pathogens. 2019;15(9):e1008067. Epub 2019/09/13. doi: 10.1371/journal.ppat.1008067. PubMed PMID: 31513651.

101. Thomann B. Gouverner les populations laborieuses dans le Japon impérial : vulnérabilité, biopolitique et citoyenneté (1868-1945). Histoire Institut d’Etude Politiques de Paris. 2012;https://hal.archives-ouvertes.fr/tel-03222520.

102. Shimao T. [Tuberculosis and its control--lessons from the past and future prospect]. Kekkaku : [Tuberculosis]. 2005;80(6):481–9. Epub 2005/09/01. PubMed PMID: 16130906.

103. Aoki K. [Epidemic of tuberculosis in Meiji and Taisho eras in Japan and excess deaths from tuberculosis in females]. Kekkaku : [Tuberculosis]. 1995;70(8):483–90. Epub 1995/08/01. PubMed PMID: 7564060.

104. Anonymous. History of Silk Wikipedia [cited 2021 2021, June 9th]. Available from: https://en.wikipedia.org/wiki/History_of_silk.

105. Wikipedia. Ashio Copper mine https://en.wikipedia.org/wiki/Ashio_Copper_Mine: Wikipedia; 2021 [cited 2021 May 31st 2021]. Available from: https://en.wikipedia.org/wiki/Ashio_Copper_Mine.

106. Ota M, Hoshino Y, Hirao S. Analysis of 605 tuberculosis outbreaks in Japan, 1993–2015: time, place and transmission site. Epidemiology and infection. 2021;149:e85. Epub 2021/03/22. doi: 10.1017/S0950268821000625.

107. Sola C, Ferdinand S, Sechi LA, Zanetti S, Martial D, Mammina C, et al. Mycobacterium tuberculosis molecular evolution in Western Mediterranean Islands of Sicily and Sardinia. Infect Genet Evol. 2005;5(2):145–56.

